# COVID-19 incidence trends between April and June 2020: A global analysis

**DOI:** 10.1101/2020.07.07.20148007

**Authors:** Vincent Amanor-Boadu, Kara Ross

**Affiliations:** Department of Agricultural Economics, Kansas State University, Manhattan, Kansas, USA

**Author notes:** These authors contributed equally to this work.

## Abstract

The study sought to investigate how the number of confirmed cases of COVID-19 have evolved in the most recent three months across the world, and what insights the trends may provide about the second half of the pandemic’s first year using a situation analysis approach based on national income, temperature, trade intensity with China, and location defined by longitude and latitude. The study confirmed the negative relationship between COVID-19 cases and temperature. It contributed to the resolution of the conflicting results about latitude after organizing it into a categorical variable instead of its continuous form. This approach works because the average temperature in the 15°S to 15°N region remains similar to the average temperatures in both the Above 15°N region and the Below 15°S region during their summer months because the 15°S to 15°N region does not experience the marked seasonal changes in temperature. Given the negative association between temperature and case numbers, this suggests that countries in the 15°S to 15°N region might continue exhibiting the low numbers they have thus far exhibited through the second half of this year, even as numbers climb in the Below 15°S region. To succeed, their policymakers must control importation of the disease by implementing effective testing, quarantining, and contact tracing for people entering their borders. Policymakers in countries Below 15°S region may manage their inherent risks by applying lessons learned from countries in the Above 15°N region during these past months. Such preventative measures may allow the world to avoid the drastic lockdown policies and facilitate rapid global economic recovery from this pandemic.

## Introduction

Six months after the outbreak of COVID-19 in Wuhan City, China, and about three months after the World Health Organization declared it a pandemic, the disease had infected nearly 8 million people and killed more than 433 thousand globally (1). The distribution of the infections (and the deaths) has so far been disproportionally centered in Europe and North America. While Europe and North America account for about 16.6% of global population, their share of the global confirmed COVID-19 cases as of June 15, 2020 was about 57.4%. Unlike previous viral outbreaks – SARS, AIDS, Ebola, MERS, and others – this one seems to be following a different path of attacking, thus far, higher income countries more. The 2018 average gross national income (GNI) per capita of about $35,805 and $34,122 in Europe and North America, respectively, contrasts with an average global GNI per capita of approximately $11,182 (excluding North America and Europe). These distributions have led to some speculations that location and income might explain the COVID-19 incidence and death rates around the world.

The cumulative number of confirmed cases in a country is a function of the infection rate and the effectiveness of non-pharmaceutical intervention public health policies implemented to control infections, including shelter-in-place, closing of schools, non-essential shopping, recreational and social events, such as sports, festivals, weddings and funerals (2). By the end of March, more than 1.7 billion people worldwide were under some form of lockdown, and a week later, the number had increased to almost 4 billion (3). While the motivation of these lockdowns was to slow the infection rate by reducing transmissions among people (4), the extent of their economic implications were uncertain. Therefore, understanding how the disease can be managed without some of these adverse economic outcomes could be helpful to many countries, especially those with smaller economies that cannot afford social support for their citizens, as found with the stimulus and bailout packages in Europe and North America (5,6).

In an April 2020 study, Jüni et al. (7) use a prospective cohort study approach over a 14-day period as the pandemic was beginning to unfold in most countries (March 7 to March 21, 2020) to explore the association between COVID-19 growth in 144 countries and territories and income and latitude. They conclude that neither income nor latitude is statistically significant in its association with COVID-19 growth. Heneghan and Jefferson (8) explored the association between the same variables using total number of cases as of April 28, 2020 in 128 countries as the dependent variable. They concluded that both income and latitude are strongly associated with COVID-19 incidence and deaths. They go on to say that low community infections in countries with high temperature and high relative humidity may be explained by their locations. Livadiotis (9) confirmed a negative relationship between environmental temperature and infection growth rates in Italy and the United States, lending support to the latitude (as proxy for temperature) hypothesis. Another study suggested that the low mortality from COVID-19 in countries south of Latitude 35°N may be a result of Vitamin D’s role in mitigating disease severity (10), which further contribute to temperature’s role in the unfolding story. Despite the foregoing, there is still some debate about the location characteristics and the infection numbers because of the changing environment (11,12). Apart from these climatic characteristics, there has also been suggestions of the association of the disease with age (13,14). A recent Pew Research Center study (15) confirmed a correlation between the incidence of COVID-19 and the proportion of the population who are 60 years or older.

Because longitude does not directly affect temperature and relative humidity, it has so far been ignored in COVID-19 studies. Yet, the distribution of COVID-19 infections shows that there are differences as one moves away from the prime meridian. Understanding the extent to which it is associated with the prevalence of the disease could open up different exploration avenues and produce alternative insights. To this end, this study seeks to contribute to resolving some of the outstanding issues surrounding the distribution of COVID-19 using all relevant and available data on infections from the beginning of the outbreak in China to date. It hopes to provide policymakers with insights into how they might deal with the pandemic in their jurisdictions as seasons shift, with the north moving into summer and the south moving into winter (16).

## Data and methods

### Data

The study uses cross-sectional data for 211 countries and territories to explore the association between the explained variable of interest, total confirmed COVID-19 cases for each jurisdiction between December 31, 2019 and June 15, 2020, and a number of explanatory variables in line with the literature. They included income, location (specified as latitude and longitude of the country’s capital city), average temperature between December and March, and trade intensity. Using the cumulative number of cases between the onset of the pandemic and a particular end date makes this study similar to Heneghan and Jefferson (8). This approach, instead of taking only a part of the data, ensures that all adjustments made by reporting agencies to address apparent reporting irregularities at the beginning of the pandemic are captured (17,18). The case data, along with population data, were obtained from the COVID-19 web pages of the European Centre for Disease Prevention and Control (1). From these two variables, the total number of cases per one million population was developed and used as the explained variable in the study.

Income was represented by the 2018 Gross National Income (GNI) per capita, as estimated and presented by the World Bank (19). The latitude and longitude of jurisdictions’ capital cities were downloaded from Dataset Publishing Language (https://developers.google.com/public-data/docs/canonical/countries_csv), and supplemented with data from GeoHack, a client hosted by Wikipedia’s Toolforge (https://tools.wmflabs.org/geohack/), which provides links to various mapping services on Wikipedia. Age distribution data were obtained from the United Nations Department of Economic and Social Affairs (20).

Because a major source of COVID-19 is imported infections (21), trade intensity is introduced as a potential explanatory variable. Trade intensity is defined as the quotient of a focus country’s total trade (imports and exports) with a target country and its total trade with the world. China, being the source of the coronavirus, is the target country in this scenario, and the focus countries are all the other countries. Trade data for 2018 were procured for the World Bank’s World Integrated Trade Solution (WITS) (https://wits.worldbank.org/) to develop the trade intensity indicator. Finally, average temperature for December through March for capital cities was calculated from monthly temperature data obtained from Wikipedia’s list of cities by average temperature (22) and used to estimate the average temperature for December through March.

## Methods

Imported COVID-19 spread, in contrast to community spread, is through travel to specific locations and/or in transit, and from travel to countries by carriers of the virus. National income is used as a proxy for travel, both for a country’s residents and as a tourism destination. In recent years, China has been a popular destination for business travel, growing from $32 billion in 2000 to $347 billion in 2017 (23), even as Chinese international tourists to higher income countries have increased (24). Eder et al. (25) report that more than 430,000 people arrived in the United States, mainly in New York, San Francisco and Los Angeles from China between January 2020 and middle of March 2020. These cities became centers of the pandemic in the early days of the disease. Pullano et al. (26) observe that the first 41 infections of COVID-19 outside of China were all traceable to people who arrived from Wuhan to Europe, North America, Oceania and Asia. Similarly, early infections in Africa were traced to travelers coming from Europe, Asia, and South America (27). Given the foregoing, it is hypothesized that the income elasticity of confirmed infections is positive, holding all other influencing variables constant. The argument is that as trade intensity with a target country increases, the number of business travel between the countries increase. If there is an infection risk in the target country, then the risk of importing the infection increases. Trade intensity is, therefore, hypothesized to have a positive association with confirmed cases, holding all other influencing variables constant.

Because there seems to be a negative association between the virus and ambient temperature (28), it is hypothesized that the temperature elasticity of confirmed infections is negative, holding all other things constant. If this holds, then it is to be expected that the infection rates in the higher latitudes should decline as those regions enter their summer months, and they should increase as the countries in the lower latitudes enter their winter months. While most studies exploring the effect of location on COVID-19 cases have used latitude as an associative variable, none has paid attention to the other location variable, i.e., longitude. Yet, the incidence map (29) shows that there seem to be differences in the cumulative cases as one moves from east to west. It is, therefore, included in this study as an explanatory variable for cumulative confirmed cases of COVID-19. It is hypothesized, on the basis of the incidence maps, that moving east decreases the incidence of COVID-19.

The foregoing suggests that the number of confirmed cases on June 15, 2020 in each country, *C*_*i*_, was influenced by a vector of explanatory variables defined to encompass the following: (i) the average income per person for each country; (ii) the location of the country (latitude and longitude); (iii) the country’s average temperature between December and March; and (iv) its trade intensity with China. The conceptual model is structured as follows:

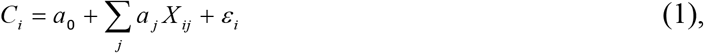

where *a*_*j*_ defines the estimated regression coefficients and *ε* is the regression error term assumed to be independently and identically distributed with a zero mean and a constant variance. The logarithm form of the number of confirmed cases is used in the regression estimation. The individual and collective effect of latitude and temperature on the number of confirmed cases are compared through multiple estimations to facilitate the best overall fit for the ordinary least square (OLS) regression model specified in Equation (1). Preliminary analysis suggested the statistical insignificant of age, and hence it was not included in the ensuing analysis. The models are estimated using Stata 16 S/E (30).

## Results and Discussion

### Summary Statistics

Total population of the 211 countries and territories included in the study was 7.56 billion, covering about 99.7% of global population estimate for June 2019. The total number of confirmed COVID-19 cases globally on June 15, 2020 was just under 7.9 million, compared to a little over 2 million on April 16, 2020, and approximately 2.9 million on May 15, 2020. Thus, June’s numbers were nearly 175% higher than May’s numbers, which was approximately 41% higher than April’s.

Table 1 presents the summary statistics for the variables used in the analyses. It shows that the average and median of total confirmed COVID-19 cases on April 16, 2020 were, respectively, about 619 and about 93 per million people. The average increased by about 70.1% a month later and by almost 165% two months later. The median, on the other hand, increased by 136% between April and May, and by more than 315% between April and June, indicating the number of cases is increasing in many countries instead of in a few countries.

**Table 1.** Summary statistics for variables used in the analysis.

| **Variable** | **N** | **Mean** | **Std. Dev.** | **Median** | **Minimum** | **Maximum** |
| --- | --- | --- | --- | --- | --- | --- |
| **Population in million** | 211 | 35.85 | 138.49 | 6.96 | 0.00 | 1,392.73 |
| **Cases/million (April 16)** | 211 | 618.97 | 1,419.38 | 93.25 | 0.00 | 11,602.78 |
| **Cases/million (May 15)** | 211 | 1,056.54 | 2,127.17 | 220.48 | 0.47 | 19,180.11 |
| **Cases/million (June 15)** | 211 | 1,640.23 | 3,160.26 | 387.39 | 0.93 | 28,616.55 |
| **GNI/capita (\$ million)** | 203 | 1.78 | 2.59 | 0.66 | 0.03 | 19.30 |
| **Latitude (Degrees)** | 211 | 20.30 | 23.95 | 18.42 | -51.80 | 71.71 |
| **Longitude (Degrees)** | 211 | 13.52 | 61.95 | 17.68 | -149.41 | 179.41 |
| **Average Temperature**<br>**(Dec-Mar) (Celsius)** | 209 | 17.14 | 10.11 | 20.83 | -10.90 | 30.00 |
| **China Trade Intensity** | 195 | 0.14 | 0.12 | 0.10 | 0.00 | 0.64 |

The top-10 jurisdictions with the highest total number of cases per million on June 15, 2020 in descending order Qatar (28,617), San Marino (20,542), Bahrain (11,614), Andorra (11,077), and Chile (9,306). The rest are Kuwait (8,682), Singapore (7,201), Peru (7,182), Luxembourg (6,697), and United States (6,401). Only half of the top-10 countries in June were also in the top-10 in April: San Marino, Andorra, Luxembourg, United States and Qatar. San Marino had moved from the jurisdiction with the highest number of cases per million in April to the second in June, and Qatar had moved from the tenth position to the top position. Luxembourg and the United States moved from the third and ninth positions, respectively, to the ninth and tenth positions. Iceland, Spain, Belgium, and Ireland had dropped out of the top-10 list by June. The coefficient of variation for the top-10 by number of cases per million for the three months decreased from 1.26 to 0.62 between April and June, suggesting that the distribution across the countries was becoming less variable.

Countries and territories were organized into three latitude categories to explore differences in their number of cases per million, average temperature between December and March, and GNI/capita. The categories are: (1) Above Latitude 15°N; (2) Below Latitude 15°S; and (3) Between Latitude 15°N and Latitude 15°S. Table 2 presents the median for the number of cases per million population, GNI/capita, and temperature by the latitude categories. Across all variables, the medians for the 15°N to 15°S category were more similar to those for the Below 15°S category. For example, the differences in the average temperature between 15°N to 15°S and Above 15°N category are about 17.8°C (p < 0.000) and between Below 15°S and Above 15°N was approximately 14°C (p < 0.000). However, the temperature difference between Below 15°S and the 15°N to 15°S categories was about 3.9°C (p < 0.000). Despite the statistically significant difference in their temperatures, there was no statistical difference between their average number of cases (123.5 cases/million, p < 0.755).

**Table 2.** Median of cases/million, average temperature, and GNI/capita by latitude categories.

| **Latitude Categories** | **April Cases** | **May Cases** | **June Cases** | **Average Temperature (°C)** | **GNI/Capita (\$/person)** |
| --- | --- | --- | --- | --- | --- |
| **15°S to 15°N** | 26.09 | 100.09 | 211.46 | 26.40 | 3,420.00 |
| **Above 15°N** | 392.84 | 806.56 | 1,146.64 | 6.25 | 17,430.00 |
| **Below 15°S** | 41.11 | 148.42 | 226.14 | 21.49 | 5,860.00 |
| **Total** | 93.25 | 220.48 | 387.39 | 20.83 | 6,610.00 |

## Regression Results and Discussion

The OLS model specified in Equation (1) was estimated using the log of the number of cases per million as the explained variable, and gross national income per capita, longitude and trade intensity, and different combinations of latitude, temperature, and latitude categories (defined above) as the explanatory variables. The combinations led to the estimation of five different models for each of the three months (April, May, and June): Model 1 involved introducing latitude alone; Model 2 was latitude and temperature; Model 3 was latitude categories alone; Model 4 introduced temperature and latitude categories; and Model 5 considered temperature alone. The model presenting the best results for each month, based on adjusted R^2^, assuming all other variables are behaving appropriately, is deemed the most qualifying in explaining total incidence. A test for multicollinearity using variable inflation factor (VIF) suggested its absence from all the estimated models, with the mean VIF below 10 in all cases. Additionally, the F-statistics for all the models were statistically significant (p < 0.000). The number of observation (countries) increased from 183 in April to 189 in May and June because six jurisdictions that had zero cases per million in April (i.e., Comoros, Hong Kong, Lesotho, Macao, Tajikistan, and Western Sahara) had positive number of cases per million in the other two months.

Model 3 produced the best regression results based on adjusted R^2^ for all three months. The results, presented in Table 3, show that the adjusted R^2^ for April as higher than for May, which was higher than for June. The decreasing coefficient of determination may be attributed to the decreasing variability in the number of COVID-19 cases per million over time, confirmed by the coefficient of variation statistics discussed above. The results show that the log of confirmed number of COVID-19 cases per million people is increasing in the GNI per capita at a decreasing rate, with both the linear and quadratic forms of GNI per capita being statistically significant (p < 0.000). Furthermore, coefficients on the linear and quadratic forms of GNI per capita both decreased in absolute terms over time. The GNI/capita elasticities of case numbers per million for April, May and June were, respectively, 0.37 (p < 0.000), 0.25 (p < 0.000), and 0.75 (p < 0.000) at the mean. The income elasticity for the three latitude categories – 15°N to 15°S, Above 15°N, and Below 15°S – for June were 0.17 (p < 0.000), 0.14 (p < 0.000), and 0.18 (p < 0.000), respectively. For May, the elasticities were 0.42 (p < 0.000), 0.32 (p < 0.000), and 0.31 (p < 0.000). And for April, the estimated elasticities were 0.27 (p < 0.000), 0.22 (p < 0.000), and 0.43 (p < 0.000). The results also suggest that being in the Above Latitude 15°N instead of in the 15°N to 15°S category is associated with an increase of 1.2% in the number of confirmed cases (p < 0.000) for May and June and about 1.3% (p < 0.000) for April. To this end, it is concluded that the hypothesis that the GNI per capita elasticity of the number of confirmed cases is positive cannot be rejected. However, because the categorical form of latitude provided a stronger explanatory power than the continuous form, the discontinuity character of latitude is recognized. The hypothesis that the latitude elasticity of the number of confirmed cases is positive is not rejected for the Above 15°N category, but rejected for the Below 15°S category, using the 15°N to 15°S category as the reference. Trade intensity was not statistically significant in explaining total case numbers in May and June, but was in April (p < 0.026) and it exhibited the wrong sign. That is, a percentage point increase in the China trade intensity in April was associated with about 2.3% decline in the total number of cases per million. The lockdown and banned travel policies that started in March in most jurisdictions may explain why the trade intensity elasticity of COVID-19 cases per million on April 16, 2020 of −0.18 was not statistically significant p < 0.499).

**Table 3.**
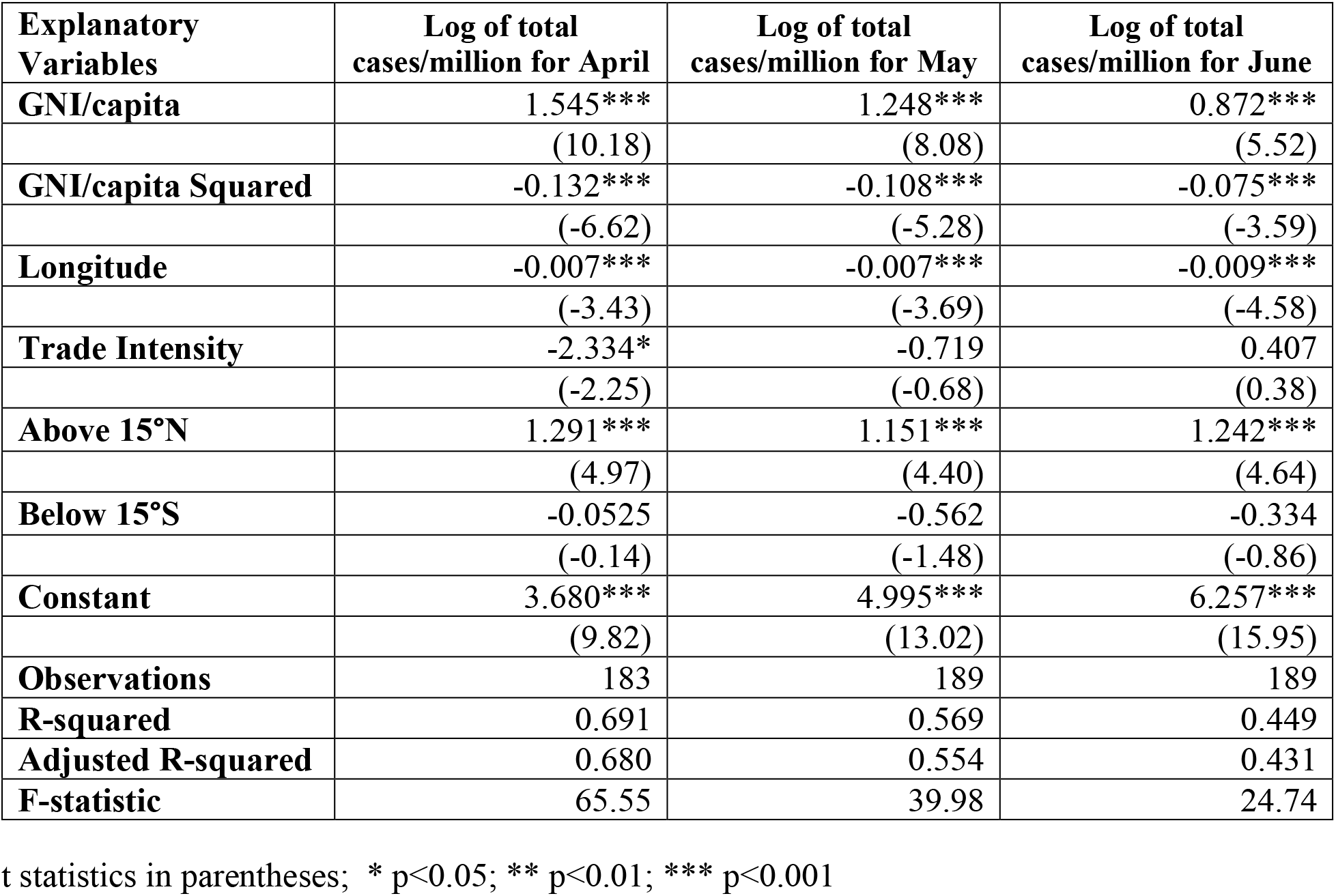
Comparing Model 3 regression results for April, May, and June cumulative number of COVID-19 cases/million.

| **Explanatory Variables** | **Log of total cases/million for April** | **Log of total cases/million for May** | **Log of total cases/million for June** |
| --- | --- | --- | --- |
| **GNI/capita** | 1.545*** | 1.248*** | 0.872*** |
|  | (10.18) | (8.08) | (5.52) |
| **GNI/capita Squared** | -0.132*** | -0.108*** | -0.075*** |
|  | (-6.62) | (-5.28) | (-3.59) |
| **Longitude** | -0.007*** | -0.007*** | -0.009*** |
|  | (-3.43) | (-3.69) | (-4.58) |
| **Trade Intensity** | -2.334* | -0.719 | 0.407 |
|  | (-2.25) | (-0.68) | (0.38) |
| **Above 15°N** | 1.291*** | 1.151*** | 1.242*** |
|  | (4.97) | (4.40) | (4.64) |
| **Below 15°S** | -0.0525 | -0.562 | -0.334 |
|  | (-0.14) | (-1.48) | (-0.86) |
| **Constant** | 3.680*** | 4.995*** | 6.257*** |
|  | (9.82) | (13.02) | (15.95) |
| **Observations** | 183 | 189 | 189 |
| **R-squared** | 0.691 | 0.569 | 0.449 |
| **Adjusted R-squared** | 0.680 | 0.554 | 0.431 |
| **F-statistic** | 65.55 | 39.98 | 24.74 |

t statistics in parentheses; \* $p < 0.05$ ; \*\* $p < 0.01$ ; \*\*\* $p < 0.001$

Temperature, while a substitute for latitude, is subject to change while latitude is not, making it an imperfect substitute. For all three months, in Model 1 and Model 5 (where the effects of latitude and temperature were independently tested), it was found that, holding all other variables constant, the coefficient for latitude was less than half of that for temperature, but both were statistically significant (p < 0.000). Additionally, the two variables exhibited their expected signs.

As indicated in the data and methods section, previous studies have ignored longitude, may be because it had nothing to do directly with temperature and humidity as latitude does. However, there was a clear trend in incidence maps showing that longitude was a factor. The results from all the estimated models indicate a statistically significant negative coefficient for longitude. For Model 3, the longitude coefficient was −0.007 (p <0.000), for April and May, and −0.009 (p < 0.000) for June. The sign suggests that increasing longitude, i.e., moving eastwards, is associated with decreasing COVID-19 number per million. The estimated longitude elasticity of cases/million for June is −0.29 (p <0.000), i.e., for each percentage increase in longitude, there is about three-tenths of a percent decrease in the number of COVID-19 cases per million population.

## Conclusions

This research has contributed to the location debate in a very substantive way. It has shown both longitude and latitude are statistically significant covariates for the log of total number of COVID-19 cases between December 31, 2019 and June 15, 2020. It also showed a significant correlation between latitude and temperature, supporting the speculations that COVID-19 cases might be following the seasons. Using GNI/capita as a proxy for travel, the results showed that while remaining statistically significant, the impact of this variable waned over time. This is expected because of the intervention policies that virtually stopped global travel by April. This is confirmed by the fact that while latitude (Model 1) and temperature (Model 5) both exhibited their hypothesized relationships with the number of confirmed cases of COVID-19, neither provided as strong a model as using latitude categories in place of both variables. As the season switches, it is expected that higher temperatures in the north would reduce its case numbers (9,31). However, this will only happen if community spread is contained. However, as is already unfolding in the United States (32) and in Germany (33), lifting the lockdown has probably increased local spread of the infection across local communities as susceptible (unexposed) people exiting their shelters-in-place encounter infected people. A few countries in the Below Latitude 15°S – Brazil, Peru, and Chile – have already started seeing spikes in their numbers. It is still too early to confirm that it is due mainly to the changing seasons, and not to changing government policies about shelter-in-place and people’s commitment to social distancing guidelines.

The countries and jurisdictions lying between Latitude 15°N and 15°S do not experience the seasonal changes the others experience, and may therefore, not see major changes in their COVID-19 numbers in the second half of the year, as long as they effectively control imported infections, and continue to manage their current infected judiciously. If they succeed at this, then they can avoid the extensive lockdowns and their attendant adverse effects on national and local economies. Yet, it is important for public health officials everywhere to remain vigilant because of the inherent possibilities for mutations of the virus (34), and the uncertainty about the effects of these mutations on infectivity (35,36). Finally, the specific implications of the negative association with the longitude variable is unclear, yet its role in defining location, and its strong statistical significance suggests that it may not be ignored. It is, therefore, recommended that future research explores this further.

## Data Availability

All data used are available publicly.

## Acknowledgments

Our gratitude to the staff at the European Centre for Disease Prevention and Control, World Health Organization, and other UN agencies who collect, process, and organize data and make them available for researchers to use in their work.

